# Geometric versus hemodynamic indexes for rupture-destined aneurysms: a retrospective cohort and a repeated-measures study

**DOI:** 10.1101/2023.02.23.23286386

**Authors:** Chan-Hyuk Lee, Hyo-Sung Kwak, Hyun-Seung Kang, Keun-Hwa Jung, Seul-Ki Jeong

## Abstract

**Background and purpose:** A proper stratification of intracranial aneurysms (IA) is critical in identifying rupture-destined aneurysms (RDA) and unruptured intracranial aneurysms (UIA). We aimed to determine the utility of geometric and hemodynamic indexes in differentiating RDA and UIA, and to examine the characteristics of natural evolutionary changes of UIA.

**Methods:** RDA was defined as having subsequent subarachnoid hemorrhage (SAH), and UIA was examined using follow-up time-of-flight magnetic resonance angiography (TOF-MRA). In addition to geometric indexes such as aspect or size ratio, aneurysmal signal intensity gradient (SIG), an *in-vivo* approximated wall shear stress (WSS) from TOF-MRA, was measured. The difference (delta) between the maximum and minimum values of SIG in an aneurysm compared to parent arterial values was designated as the delta-SIG ratio.

**Results:** This study analyzed 20 RDA in 20 patients and 45 UIA in 41 patients with follow-up TOF-MRA. While geometric indexes did not show significant differences between the RDA and UIA, the delta-SIG ratio was significantly higher in the RDA than in the UIA (1.5±0.6 vs. 1.1±0.3, P=0.032). The delta-SIG ratio showed a significantly higher area under the receiver operating characteristics curve for SAH than the size ratio (0.72 [95% confidence interval (CI), 0.58–0.87] vs. 0.56 [95% CI, 0.41–0.72], P=0.033). The longitudinal re-examination of TOF-MRA in the UIA group showed evidence of aneurysmal growth with hemodynamic stability.

**Conclusions:** The delta-SIG ratio showed significantly higher discriminatory results between RDA and UIA compared to geometric indexes. Aneurysmal rupture risk should be assessed by considering both geometric and hemodynamic information.

This study was registered on ClinicalTrials.gov (NCT05450939).

## Introduction

Incidental diagnosis of intracranial aneurysm (IA) has increased with the widespread use of neurovascular imaging techniques such as computed tomographic angiography (CTA), magnetic resonance angiography (MRA), and digital subtraction angiography (DSA). The incidence of IA ranges from 2% to 3% in the general population^1,2^ and more than 4% in patients with autosomal dominant polycystic kidney disease.^3,4^ IA should be carefully managed because it can lead to fatal subarachnoid hemorrhage (SAH), with high rates of mortality or morbidity with severe neurologic sequelae.

Approximately 1% of patients with IA experience catastrophic hemorrhage.^5-8^ The high detection rate of IA raises the question of the most appropriate method for categorizing the risk for rupture and SAH. Previous studies have used geometric indexes such as aspect ratio^9^ or size ratio^10^ and hemodynamic analysis using computer fluid dynamics (CFD)^11,12^ to differentiate between ruptured and unruptured intracranial aneurysms (UIA). However, further research is necessary to better understand the pathophysiology and natural course of evolution of IA, including identifying novel radiological markers for rupture.^13^

The previous studies that aimed to differentiate ruptured aneurysms from UIA were mostly cross-sectional studies.^14-16^ However, the validity of simple cross-sectional comparisons between ruptured aneurysms and UIA needs to be carefully evaluated considering that aneurysms evolve over time, exhibits growth,^17^ and even increase in size after rupture.^18^ Therefore, the design of studies on risk stratifications for aneurysm rupture should take into account the temporal aspect of aneurysms.

For IA, a longitudinal re-examination is needed to track the changes in geometry and hemodynamics and to better define UIA. It is possible that some ruptured aneurysms had features that indicate a high risk for rupture while in the UIA stage. At this stage, such aneurysms can be considered as rupture-destined aneurysms (RDAs). A comparison of RDA and UIA and a longitudinal re-examination of UIA would be helpful in understanding the nature of aneurysmal evolution and provide a practical perspective.

In addition to geometric indexes, hemodynamic information is useful for the proper categorization of IA. Wall shear stress (WSS) provides combined information on vascular flow and geometry, and has shown its usefulness in categorizing IAs.^19^ To measure WSS in a non-invasive manner, arterial wall signal intensity gradient (SIG) was introduced, which is an *in vivo* approximation of WSS based on an interpolation of discrete space from Time-of-Flight (TOF) MRA.^20^ The *in vivo* nature of SIG has been shown to be useful in identifying the relationship between WSS and the progression of moyamoya disease,^21^ and in determining the laterality of ischemic stroke related to large artery atherosclerosis.^22^

In the present study, we aimed to evaluate the utility of geometric or hemodynamic indexes in differentiating RDA and UIA, and to understand the characteristics of natural evolutionary changes of UIA. For this purpose, we performed (1) a retrospective cohort study among patients with IA for comparing RDA and UIA, and (2) a longitudinal re-examination of UIA. We used TOF-MRA and cerebral arterial SIG to measure geometric indexes and hemodynamic indexes.

## Methods

### Study design

This study is a single-center, retrospective cohort study that analyzed patients diagnosed with IA. We selected patients diagnosed with IA by TOF-MRA between January 2010 and December 2021. The patients were categorized into two groups according to the presence of subsequent SAH due to aneurysmal rupture. The aneurysms in patients with subsequent SAH were designated as RDA while those in patients without SAH were designated as UIA, if re-examinations of TOF-MRA were carried out. This study was approved by the institutional review board of Jeonbuk National University Hospital (CUH 2021-11-045-001) and was registered on ClinicalTrials.gov (NCT05450939). The institutional review board confirmed that this study was conducted based on appropriate guidelines and regulations (Human Research Protection Program, version 2.0).

### Selection of study subjects

#### Inclusion criteria

We included patients with RDA who fulfilled the following inclusion criteria: (1) diagnosis of a saccular aneurysm in TOF-MRA before an occurrence of aneurysmal SAH during the study period; (2) visited the hospital with clinical symptoms of headache, nuchal rigidity, focal neurologic signs, or changes of consciousness showing Hunt and Hess Grade I or higher, and SAH was confirmed on neuroimaging (brain CT or MRI); (3) SAH was caused by the previously-diagnosed saccular aneurysm, confirmed by neurovascular imaging (including CTA, MRA, or DSA) within 24 hours of the aneurysmal rupture; (4) 18 years of age or older.

For patients with UIA, the inclusion criteria were as follows: (1) diagnosis of a saccular aneurysm in the first TOF-MRA during the study period; (2) second TOF-MRA was performed before the case confirmation; (3) confirmed to be free of SAH through electronic medical chart review until the designated time (December 2021); (4) 18 years of age or older.

#### Exclusion criteria

Patients were excluded if they had: (1) any fusiform, traumatic, or mycotic aneurysms; (2) underwent endovascular or neurosurgical procedures for IA; (3) aneurysmal SAH that was clinically suspected but could not undergo imaging tests due to the patient’s medical conditions; (4) lost to follow-up or did not undergo the second TOF-MRA (in cases of UIA); or (5) intracranial aneurysms in the segment distal to M2 segment of the middle cerebral artery (MCA). TOF-MRA is based on flow-related enhancement (FRE) to build up vascular images. In the M2 segment, vascular images and signal intensities were known to be affected by in-plane saturation due to the normally horizontal course of the M1 segment of MCA.^23^

### Clinical data collection

The following information was collected by reviewing the patients’ electronic medical records: sex, age, cardiovascular risk factors (e.g., hypertension, type 2 diabetes, dyslipidemia, atrial fibrillation, smoking status), medication, laboratory findings (e.g., complete blood count, coagulation factors, liver function test, lipid profiles, and thyroid function test), and clinical prognosis (hospitalization period, and patient’s status at discharge).

### Aneurysmal indexes

#### Geometric aneurysmal indexes

We measured the maximal height and width, perpendicular height, neck diameter, and parent artery diameters of all IAs. Using these values, the following two-dimensional factors were calculated: aspect ratio,^16^ size ratio,^14^ bottleneck factor,^24^ height-width ratio,^24^ aneurysmal angle,^14^ and inflow angle^25^ (Supplementary Figure 1).

### SIG aneurysmal indexes from TOF-MRA

#### Protocol of TOF-MRA

We used a 3.0T MR machine with 32 channel cardiac coil (Philips Medical Systems, Achieva, The Netherlands) to obtain three-dimensional intracranial TOF images. The parameters of TOF-MRA were as follows: TR = 23.0 ms, TE = 3.5 ms, Resolution = 300 × 220, FOV (field of view) = 180 mm^2^, 160 slices, FA (flip angle) = 20.0°, scan duration = approximately 1 minute 53 seconds.

### Measurement of SIG aneurysmal indexes

The arterial wall SIGs, which are *in-vivo* approximated WSS based on an interpolation of discrete space from TOF-MRA, were calculated and measured with a semi-automated software (VINT, Mediimg, Inc., Seoul, Republic of Korea) according to previously reported protocol.^20^ Briefly, the signal intensities (SIs) at the iso-point (*Φ*_*a*_; *SI at position A* [Xa] *along the arterial contour line*) and at the inner point (*Φ*_*b*_; *SI at position B* [Xb]) were calculated by using a trilinear interpolation algorithm based on the positions and SIs in the eight neighboring voxels. The SIs of TOF-MRA were normalized to eliminate the offset and scale effects across the MRA datasets of participants. For each iso-point (position A), the SIG was calculated from the difference in SIs between points A and B as follows:

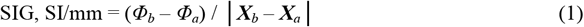

As aneurysmal indexes, SIG mean, standard deviation (SD), and delta (Δ, difference between maximum and minimum SIG) ratios comparing an aneurysm and its proximal two or more parent vessel segments were calculated as follows (Figure 1).

**Figure 1.**
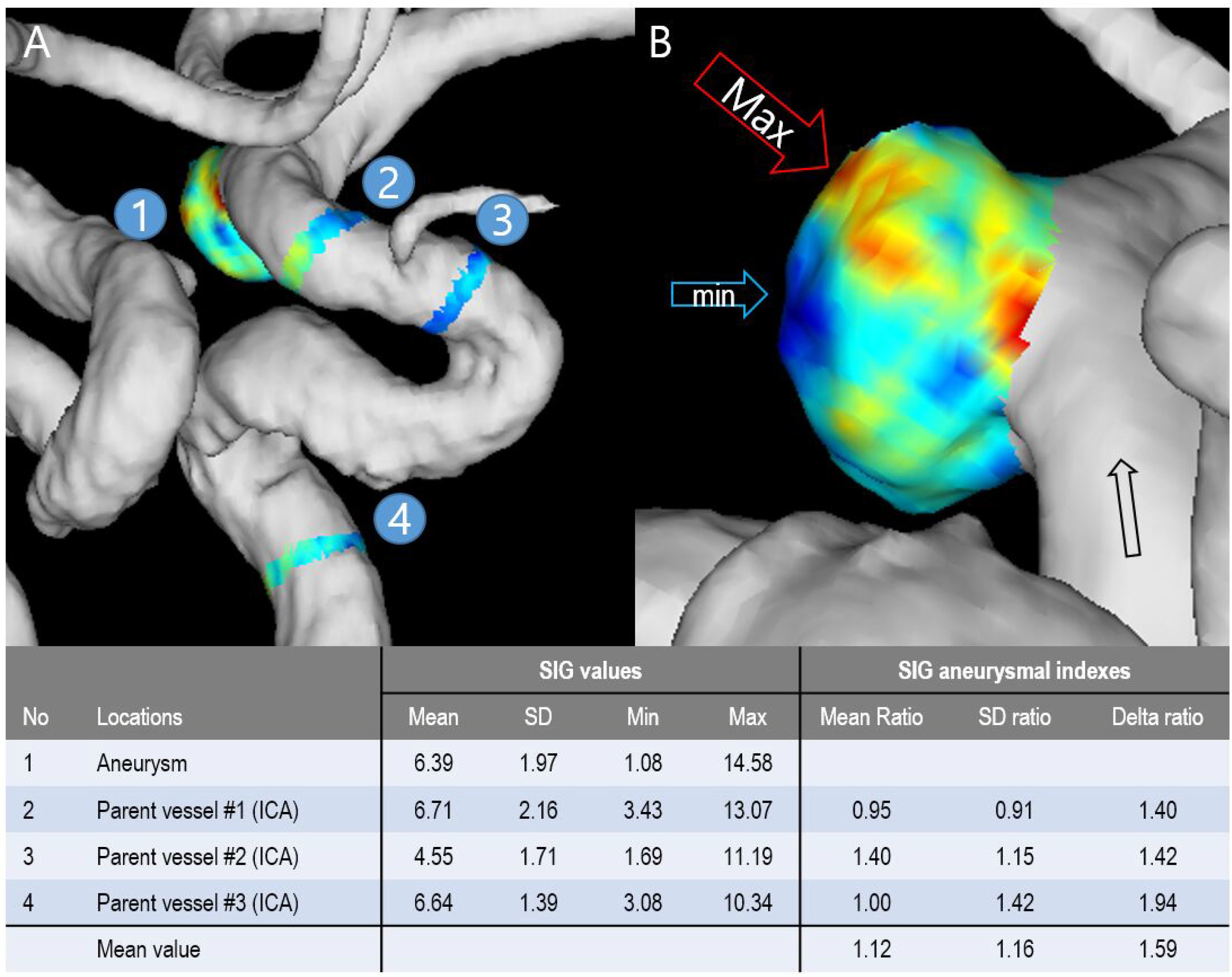
Calculations of signal intensity gradient (SIG) aneurysmal indexes. (A) A rupture-destined aneurysm (RDA) (number 1) and its parent arterial segments for reference (2, 3, and 4). (B) Detailed view of intracranial aneurysm with SIG display. Red (hot) spots are observed along the presumed flow-in line, and blue (cold) spots are observed along the outer wall, suggesting high and low shear stress zones, respectively. SIG values and their aneurysmal indexes are presented in the table. SIGs measured with a semi-automated software (VINT, Mediimg, Inc., Seoul, Republic of Korea). SD: standard deviation

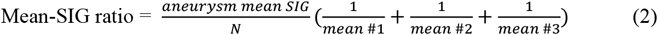

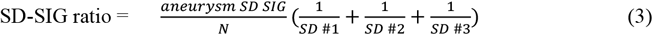

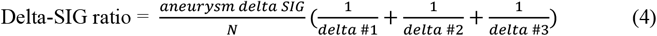

Mean #1 indicates arterial SIG mean values of the first near-parent vessel and means #2 or 3 indicate the SIG mean values of the second or third far-parent vessels. The level of mean #1 was chosen within 10 mm from the opening of the aneurysm proximally and the levels of #2 or 3 were beyond the level, as used in previous studies.^12,26^ Three proximal segments were usually used to calculate the ratios (Figure 1A); however, in the anterior communicating artery (AComA) aneurysms between two anterior cerebral arteries (ACAs), two proximal sites in each ACA (four total) were measured.

### Statistical analysis

Descriptive data for the clinical characteristics and laboratory findings of the participants are expressed as mean ± standard deviation (SD) or percentages. The Kolmogorov-Smirnov test was performed to determine distributional adequacy. Student’s *t*-test or chi-squared test was used to assess differences between groups. Three independent examiners assessed the stability and test-retest reliability among the study patients. Logistic regression analysis was performed to determine the independent association between the aneurysmal geometric or SIG indexes and the risk of SAH while adjusting for possible confounders. Receiver operating characteristic (ROC) curves were used to determine the validity of the geometric and SIG aneurysmal indexes. The areas under the ROC curves (AUC) were calculated and described with standard errors (SE) using the trapezoidal rule,^27^ and comparisons between AUCs were made using an algorithm with an estimated covariance matrix.^28^ The optimal cut-off points were chosen to balance the false-negative and false-positive rates. Statistical analyses were conducted using SPSS version 20 (IBM Corp., Armonk, NY, USA) and Stata 17 software (Stat, College Station, TX, USA).

## Results

A total of 85 IAs were initially selected, of which 24 were RDAs and 61 were UIAs. One patient with UIA and diffuse dolichoectasia was excluded because aneurysmal indexes could not be measured appropriately. IAs in the M2 segment of MCA were found in 15 patients (25.0%) in the UIA group and 4 patients (16.7%) in the RDA group (P=0.410). Two UIAs were observed in the RDA group but not followed up for the second TOF-MRA. Finally, 20 RDAs in 20 patients with subsequent aneurysmal SAH and 45 UIAs in 41 patients with follow-up TOF-MRA were included in the analysis.

Representative cases from both groups are presented in Figure 2 (A and B: RDA in AComA and internal carotid artery posterior communicating region; C and D: UIA in ACA A2 portion and AcomA). The RDA in Figure 2A showed an aspect ratio of 1.4 and a delta-SIG ratio of 1.3, while the RDA in Figure 2B showed an aspect ratio of 1.1 and a delta-SIG ratio of 1.6. The UIA in Figures 2C and 2D were re-examined 1,820 and 3,779 days after their initial assessment. The UIA in Figure 2C showed a geometric growth (aspect ratio from 2.0 to 2.7), but a decrease in hemodynamic (delta-SIG ratio from 2.0 to 1.1). The UIA in Figure 2D showed both geometric growth (aspect ratio from 1.9 to 3.1) and an increase in hemodynamic (delta-SIG ratio from 0.9 to 1.1). The locations of intracranial aneurysms are presented in Supplementary Table 1. RDAs were most commonly observed in the left internal carotid artery (40.0%), while UIAs were most commonly observed in the right internal carotid artery (26.7%).

**Figure 2.**
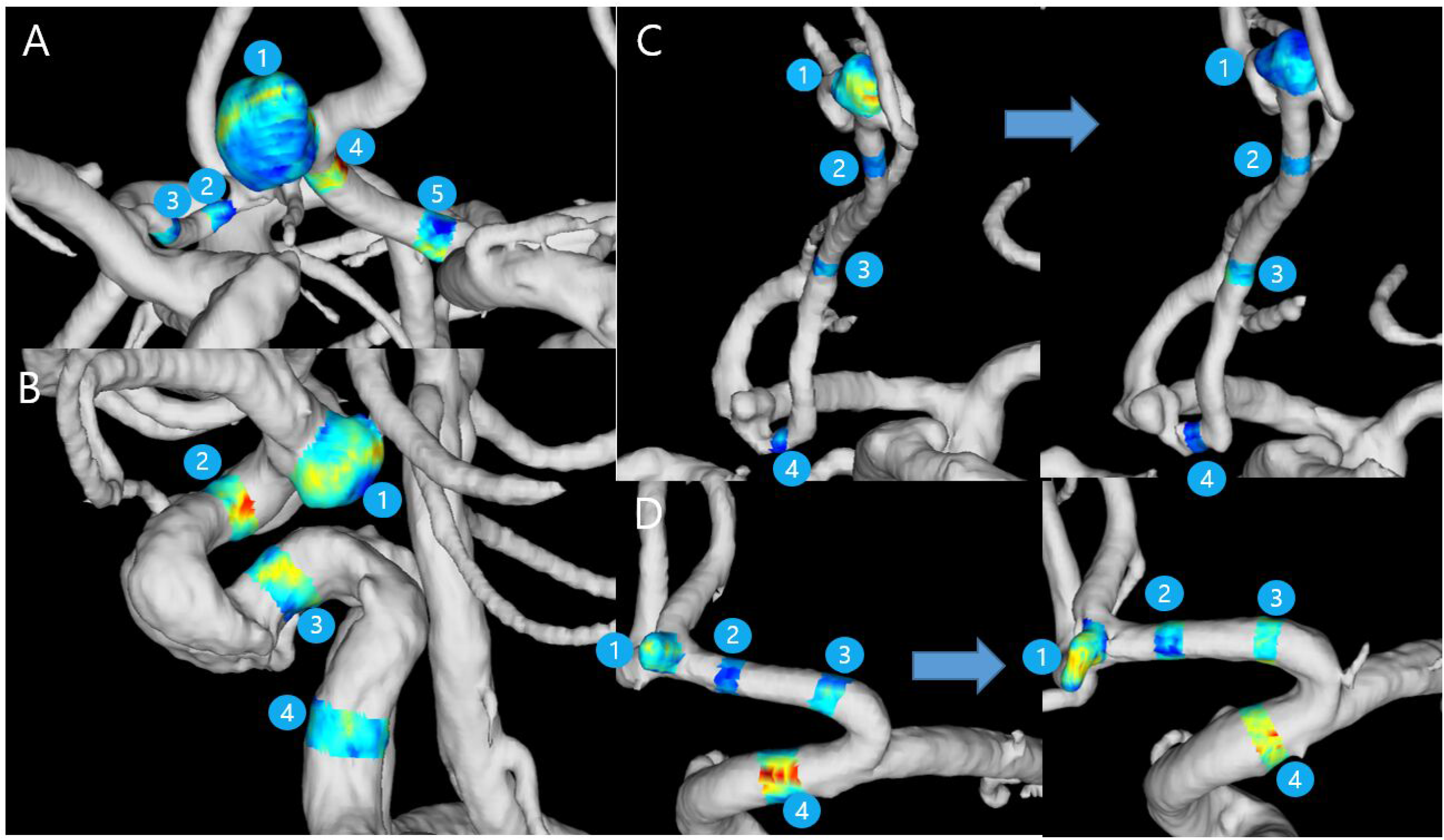
Representative cases of rupture-destined aneurysms (RDAs) and unruptured intracranial aneurysms (UIAs). Arterial SIG is displayed on the aneurysm (number 1) and its parent vessels (2, 3, 4, and 5). (A) RDA in the anterior communicating artery (maximum height: 7.9 mm, aspect ratio: 1.4, delta-SIG ratio 1.3). SAH occurred 1,539 days after the MRA exam. (B) RDA in the internal carotid artery posterior communicating region (maximum height: 4.7 mm, aspect ratio: 1.1, delta-SIG ratio: 1.6). SAH occurred 2,651 days later. (C) UIA in the A2 segment of the anterior cerebral artery. A follow-up MRA (right) was performed 1,820 days later (maximum height: 5.1 mm to 5.3 mm, aspect ratio: 2.2 to 2.7, delta-SIG ratio: 2.0 to 1.1). (D) UIA in the anterior communicating artery. A follow-up MRA was performed 3,779 days later (maximum height: 2.9 mm to 3.9 mm, aspect ratio: 1.9 to 3.1, delta-SIG ratio: 0.9 to 1.1).

The mean interval between TOF-MRA examination and SAH was 4.8 ± 2.9 years and the mean interval between the first TOF-MRA and disease-free confirmation was 12.2 ± 3.9 years (P<0.001) (Supplementary Table 2). The mean interval between the first and second TOF-MRA in the UIA group was 5.5 ± 3.4 years. While IAs were observed more commonly in women, there was no significant difference in the proportions of women between the two groups.

As for geometric indexes, there was no significant difference between the RDA and UIA groups, even considering either the first or second TOF-MRA examinations in the UIA group (Table 1). Among the SIG aneurysmal indexes, the delta-SIG ratio was significantly higher in the RDA group than in the UIA group for the first TOF-MRA (1.5±0.6 vs. 1.1±0.3, P=0.011), and the difference was increased for the second TOF-MRA (1.5±0.6 versus 1.0±0.3, P=0.002). Similar trends were observed for the other SIG indexes. In the UIA group, geometric indexes such as maximum height, perpendicular height, size ratio, and aspect ratio were significantly higher in the second examination than in the first examination.

**Table 1.** Geometric and signal intensity gradient (SIG) aneurysmal indexes

| Indexes | RDA | UIA |  |
| --- | --- | --- | --- |
|  |  | First TOF-MRA | Second TOF-MRA |
| *Geometric indexes* |  |  |  |
| Maximum height, mm | 4.0±1.7 | 3.7±2.5 | 4.1±3.2<sup>††</sup> |
| Maximum width, mm | 4.2±2.0 | 3.8±1.5 | 4.0±1.8 |
| Neck diameter, mm | 3.6±1.8 | 3.2±1.2 | 3.2±1.2 |
| Perpendicular height, mm | 3.5±1.6 | 3.2±2.2 | 3.6±2.9<sup>†</sup> |
| Size ratio | 1.4±0.8 | 1.3±1.1 | 1.6±1.3<sup>††</sup> |
| Aspect ratio | 1.1±0.5 | 1.1±0.9 | 1.3±1.1<sup>††</sup> |
| Bottleneck factor | 1.2±0.4 | 1.3±0.6 | 1.3±0.7 |
| Height width ratio | 0.9±0.3 | 0.8±0.3 | 0.9±0.4 |
| Aneurysm angle, ° | 77.0±21.1 | 81.6±21.2 | 82.4±20.9 |
| Flow angle, ° | 100.3±38.3 | 103.9±38.5 | 102.1±35.1 |
| *SIG aneurysmal indexes* |  |  |  |
| Mean-SIG ratio | 1.0±0.2 | 1.0±0.3 | 0.8±0.2<sup>**,††</sup> |
| SD-SIG ratio | 1.2±0.4 | 1.0±0.4 | 0.9±0.3<sup>**,†</sup> |
| Delta-SIG ratio | 1.5±0.6 | 1.1±0.3<sup>*</sup> | 1.0±0.3<sup>**,†</sup> |

Values are mean±standard deviation.
\*P <0.05, \*\*P <0.01 compared to the RDA
†P <0.05, ††P <0.01 between the first and second TOF-MRA examinations

Multivariate associations of geometric and SIG aneurysmal indexes for aneurysmal SAH are presented in Table 2. None of the geometric indexes based on the first TOF-MRA in the UIA group had a significant association with aneurysmal SAH; results for the geometric indexes based on the second TOF-MRA are presented separately in Supplementary Table 3. Among the SIG indexes from the first MRA, the delta-SIG ratio had an odds ratio (OR) of 1.29 (95% confidence interval (CI), 1.03–1.60, P=0.024); in contrast, all SIG indexes from the second MRA showed significant associations with aneurysmal SAH, with an OR of 1.62 (95% CI, 1.11–2.34, P=0.011) for the delta-SIG ratio, after adjustment for age, sex, free T4, and white blood cell count.

**Table 2.** Adjusted odds ratios (aORs) and 95% confidence intervals (CIs) for subarachnoid hemorrhage (SAH)

| **Indexes** | **aOR** | **95% CIs** | **P value** |
| --- | --- | --- | --- |
| *Geometric indexes, based on the first TOF-MRA* |  |  |  |
| Maximum height | 1.02 | 0.71–1.45 | 0.933 |
| Maximum width | 0.93 | 0.62–1.42 | 0.751 |
| Neck diameter | 1.15 | 0.69–1.90 | 0.591 |
| Perpendicular height | 1.08 | 0.70–1.65 | 0.732 |
| Size ratio | 0.63 | 0.21–1.86 | 0.401 |
| Aspect ratio | 1.04 | 0.36–3.03 | 0.945 |
| Bottleneck factor | 0.51 | 0.12–2.27 | 0.379 |
| Height width ratio | 4.19 | 0.32–55.23 | 0.276 |
| Aneurysm angle | 0.98 | 0.95–1.02 | 0.334 |
| Flow angle | 0.98 | 0.96–1.01 | 0.195 |
| *SIG aneurysmal indexes, based on the first TOF-MRA, per 0.1unit increase* |  |  |  |
| Mean-SIG ratio | 1.49 | 0.97–2.30 | 0.069 |
| SD-SIG ratio | 1.19 | 0.94–1.49 | 0.142 |
| Delta-SIG ratio | 1.29 | 1.03–1.60 | 0.024 |
| *SIG aneurysmal indexes, based on the second TOF-MRA, per 0.1unit increase* |  |  |  |
| Mean-SIG ratio | 1.58 | 1.04–2.40 | 0.031 |
| SD-SIG ratio | 1.64 | 1.11–2.42 | 0.013 |
| Delta-SIG ratio | 1.62 | 1.11–2.34 | 0.011 |

Adjusted for age, sex, free T4, and white blood cell count.

The performances of geometric and SIG aneurysmal indexes with AUC analysis for aneurysmal SAH are presented in Table 3 and Figure 3. Among the geometric indexes from the first exam in the UIA group, the AUC values of maximum height and aspect ratio were 0.62 (95% CI, 0.47–0.78) and 0.60 (95% CI, 0.45–0.74), respectively. In the UIA group, the geometric indexes generally showed lower AUC values with the second examination compared to the first examination. Conversely, the delta-SIG ratio from the first examination had an AUC value of 0.72 (95% CI, 0.58–0.87), which was significantly higher than those of size ratio or maximum width. From the second examination, the mean- and delta-SIG ratios had AUC values of 0.77 (95% CI, 0.65–0.89) and 0.80 (95% CI, 0.66–0.93), respectively, which were significantly higher values than those from the geometric indexes. At a cut-off point of 1.16, the sensitivity, specificity, positive likelihood ratio (LR), and negative LR of the delta-SIG ratio were 0.79, 0.66, 2.3, and 0.3 from the first examination, respectively, and 0.80, 0.73, 2.9, and 0.3 from the second examination, respectively.

**Table 3.**
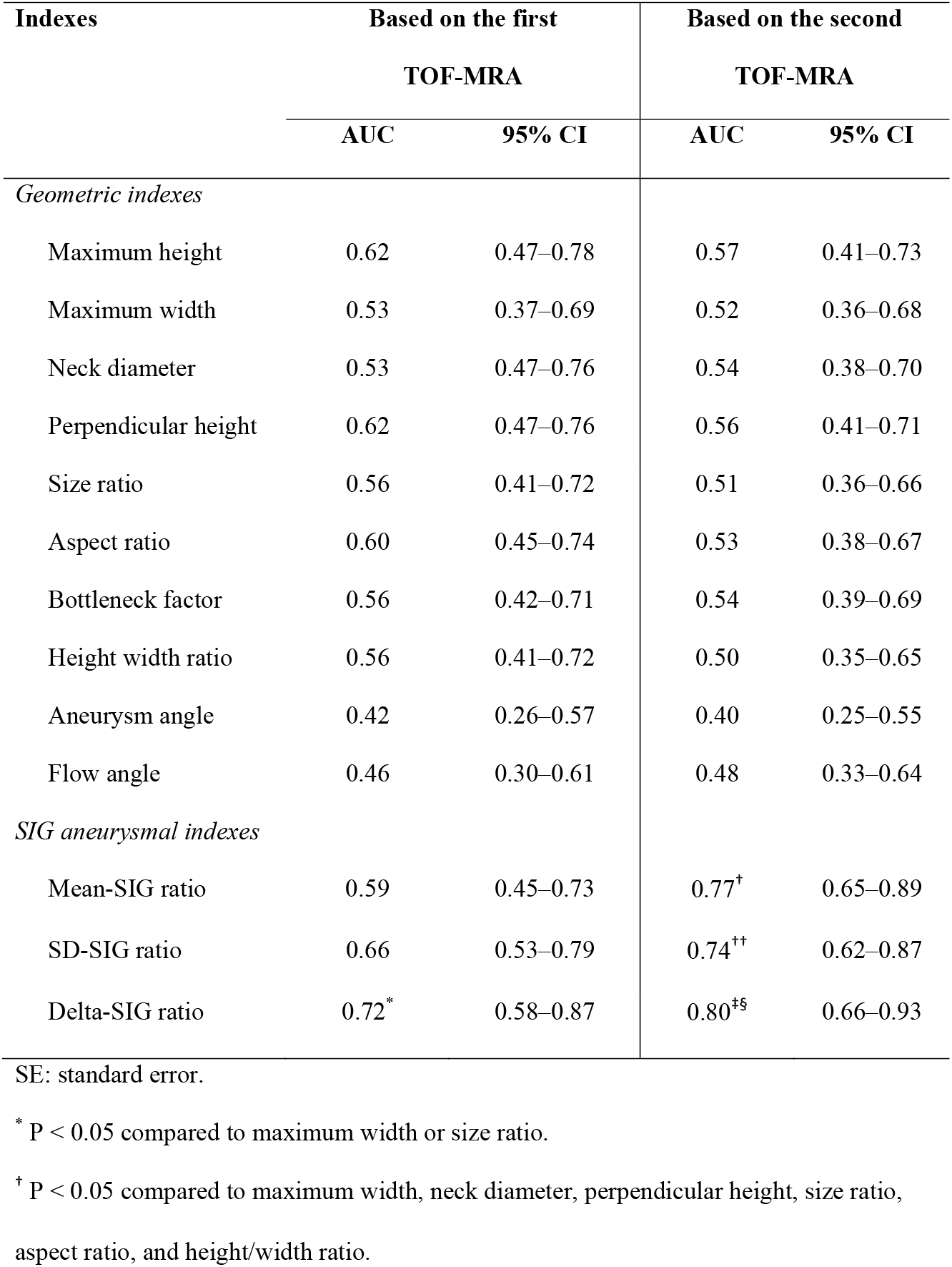
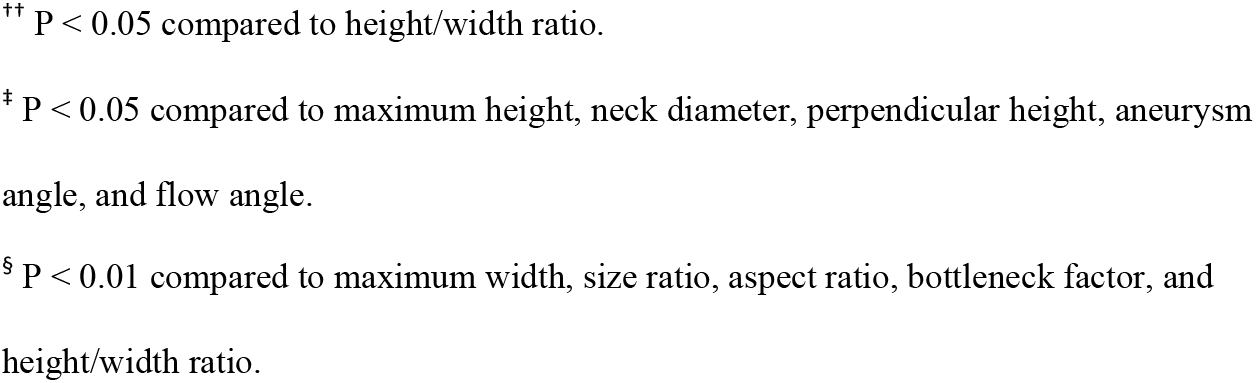
Area under receiver operating characteristic curve (AUC) analysis for aneurysmal rupture SAH according to geometric and SIG aneurysmal indexes

**Figure 3.**
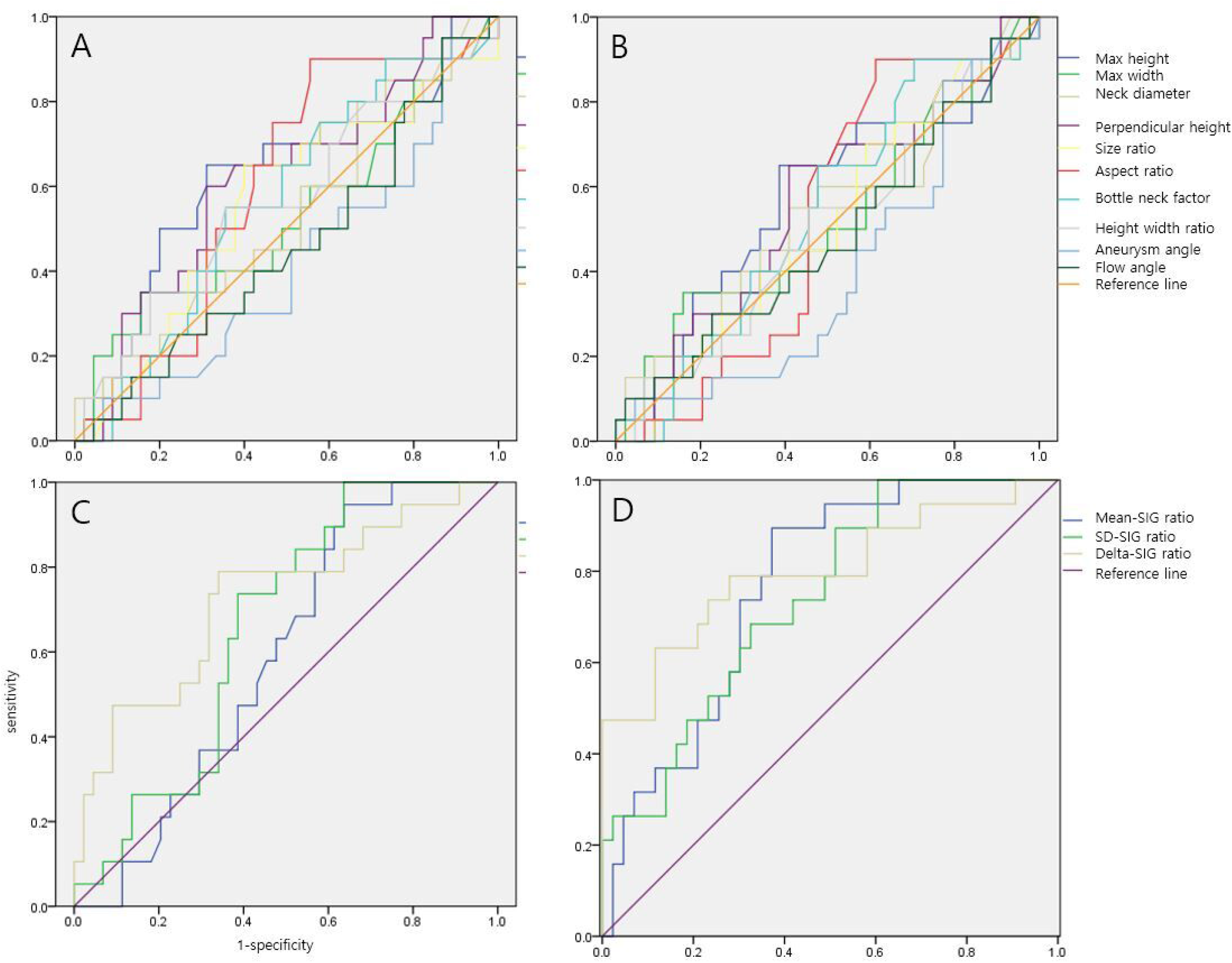
Area under the receiver operating characteristic curve (AUC) analyses of geometric and SIG aneurysmal indexes for SAH. (A and B) Geometric aneurysmal indexes based on the first TOF-MRA (A) and the follow-up TOF-MRA (B). (C and D). SIG aneurysmal indexes based on the first TOF-MRA (C) and the follow-up TOF-MRA (D).

Cross-correlations between the geometric and SIG aneurysmal indexes from the first TOF-MRA were performed to assess the construct validity, and the results are presented in Supplementary Table 4. Geometric indexes showed highly significant correlations according to the anatomic features. The aspect ratio showed significant correlations with the bottleneck factor (r = 0.784, P<0.001), height/width ratio (r = 0.721, P<0.001), and flow angle (r = 0.318, P value=0.010). In terms of the SIG indexes, the delta-SIG ratio showed a significant negative correlation with aneurysm angle (r = -0.292, P=0.020) among the geometric indexes.

The inter-rater reliability for SIG aneurysmal indexes was examined through a test-retest among three independent examiners, and the results are presented in Supplementary Table 5. All correlation coefficients were greater than 0.85 (all P < 0.001).

## 4. Discussion

In the present retrospective cohort study, we defined rupture-destined aneurysms (RDA) and compared them with unruptured intracranial aneurysms (UIA), for whom brain TOF-MRA was examined twice. We found that there was a significant difference in the hemodynamic features between the RDA group and the UIA group, and that the UIA group showed significant geometric and hemodynamic changes even within a short time span. Our key findings are as follows: (1) the RDA group showed a significantly higher delta-SIG ratio than the UIA group before aneurysmal rupture; (2) the difference in the SIG indexes between the RDA and UIA groups became more prominent in the second TOF-MRA than in the first examination, while a similar trend was not observed for the geometric indexes; (3) aneurysmal rupture risk should be assessed using both geometric and hemodynamic information.

Intracranial arteries, which are muscular arteries with no external elastic lamina and a paucity of elastic fibers in the media, are considered vulnerable to aneurysm formation or rupture.^29^ A meta-analysis reported that ruptured aneurysms have a significantly higher rate of low WSS (0-1 Pa).^11^ A case study reported that the rupture point was accompanied by markedly low WSS and high oscillating index without flow impingement.^30^ While some CFD studies reported a close relationship between high WSS regions and aneurysmal rupture,^12^ others reported that both high and low WSS regions play a role in aneurysmal growth and rupture.^16,19,26^

The delta-SIG ratio is the difference between the maximum and minimum values of SIG in the selected region. The delta-SIG ratio of an aneurysm is determined by dividing the aneurysmal delta-SIG values by the SIG values of the parent artery. A delta-SIG ratio greater than 1 indicates that the aneurysm is exposed to both higher maximal and lower minimal values of SIG compared to the parent artery. The negative correlation between the delta-SIG ratio and aneurysm angle observed in the present study suggests that aneurysms with larger delta-SIG ratios might experience enlargement in the opposite direction to blood flow, where the WSS is relatively low. The combination of strong blood flow (impetus) into the aneurysm and recirculating eddy currents on the opposite side may increase the risk of rupture.

We performed re-examinations of TOF-MRA to check aneurysmal growth or morphological changes in accordance with the current guideline.^31^ The geometric indexes from the second TOF-MRA showed decreases in the AUCs for aneurysmal SAH compared with those from the first TOF-MRA. However, aneurysmal growth was discordant with hemodynamic information in the present study. The mean- or SD-SIG ratios as well as the delta-SIG ratio based on the second follow-up MRA showed significantly higher AUCs than geometric indexes.

The two representative cases of UIA (Figures 2C and 2D) showed there could be discordant findings between growth and hemodynamics. While the aneurysm in Figure 2D demonstrated both aneurysmal growth and hemodynamic instability with delta-SIG increase, the aneurysm in Figure 2C showed aneurysmal growth but hemodynamic stability with a decrease in delta-SIG. While the case in Figure 2D would be assigned an increased risk for rupture in the follow-up examination, the case in Figure 2C should be considered carefully for potential risks. Although we could not clearly stratify the risk of these two cases, they highlight the potential for a type 1 error in risk estimation if only growth or geometric indexes are considered.

The occurrence of aneurysmal rupture is about 1.0% in patients with IA, and the prevalence of IA is ∼3% in the general population.^13^ The discrepancy between the high percentage of UIA and the low rate of rupture is not well understood, especially when considering only geometric indexes. However, this study suggests that hemodynamic stability also plays a significant role in aneurysmal rupture, even amidst growth. The relationship between hemodynamic stability and the risk of aneurysmal rupture is supported by the fact that cerebral blood flow (CBF) decreases with age^32^ and in patients with aneurysmal SAH.^33^

In the UIA group, the geometric characteristics showed significantly greater values in the second follow-up MRA than the first examination, such as maximum height, aspect ratio, and size ratio, which could be considered as indicating a heightened risk for aneurysmal rupture.^5,34,35^ The less clear differentiation between RDA and UIA in the follow-up examination seems logical because the geometric indexes of the UIA group increased over time, approaching the values of the RDA group (Table 1). This raises the question of whether the aneurysms in the UIA group are becoming increasingly vulnerable to rupture with time. Also, the presence of unruptured aneurysms more than 12 years after the first MRA raises the question of the interpretation of aneurysmal growth and its association with rupture risk.^35^ The findings of the present study suggest that aneurysmal growth or geometric indexes should be interpreted in conjunction with hemodynamic information in order to gain a better understanding of the rupture risk.

The present study has some important limitations. First, selection bias may have been present because the present study only included patients who underwent TOF-MRA examinations. Second, some patients with presumed high-risk aneurysms might have undergone endovascular therapy or neurosurgical coiling before aneurysmal SAH and would have been excluded from the present study. Third, as geometric indexes in the RDA group were measured at the unruptured stage, it is unknown if there are any differences between RDA and ruptured aneurysms. Fourth, aneurysms in the M2 segment of MCA were excluded because of technical issues. A more detailed analysis of the changes in signal intensity distal to the M1 segment is needed. Last, it is unclear whether there were concurrent changes in the risk factors for aneurysmal rupture such as normalization of blood pressure and cessation of smoking or alcohol consumption during the longitudinal re-examination of TOF-MRA.

## Data Availability

Our research data can be provided upon reasonable request.

## Conclusions

The delta-SIG ratio was significantly higher in the RDA group than in the UIA group. The diagnostic performance of the delta-SIG ratio in terms of the AUC value was significantly superior to those of geometric indexes such as aspect ratio or size ratio. The risk of aneurysmal rupture should be carefully estimated by considering the possible discrepancy between geometric and hemodynamic information. Further studies using larger patient populations and diverse ethnicities are needed to determine whether this finding is repeatable and generalizable.

## Acknowledgements

We give special thanks to Youngdo Na and Wooseok Jeong for data acquisition.

## Supplementary materials

1 Supplementary figure and 5 supplementary tables

## References

1. Vlak MH, Algra A, Brandenburg R, Rinkel GJ. Prevalence of unruptured intracranial aneurysms, with emphasis on sex, age, comorbidity, country, and time period: a systematic review and meta-analysis. Lancet Neurol. 2011;10:626–636. doi: 10.1016/s1474-4422(11)70109-0

2. Cras TY, Bos D, Ikram MA, Vergouwen MDI, Dippel DWJ, Voortman T, Adams HHH, Vernooij MW, Roozenbeek B. Determinants of the Presence and Size of Intracranial Aneurysms in the General Population: The Rotterdam Study. Stroke. 2020;51:2103–2110. doi: 10.1161/STROKEAHA.120.029296

3. Lee CH, Ahn C, Ryu H, Kang HS, Jeong SK, Jung KH. Clinical Factors Associated with the Risk of Intracranial Aneurysm Rupture in Autosomal Dominant Polycystic Kidney Disease. Cerebrovascular Diseases. 2021;50:339–346. doi: 10.1159/000513709

4. Chapman AB, Rubinstein D, Hughes R, Stears JC, Earnest MP, Johnson AM, Gabow PA, Kaehny WD. Intracranial aneurysms in autosomal dominant polycystic kidney disease. N Engl J Med. 1992;327:916–920. doi: 10.1056/NEJM199209243271303

5. Wiebers DO, Whisnant JP, Huston J, 3rd, Meissner I, Brown RD, Jr., Piepgras DG, Forbes GS, Thielen K, Nichols D, O’Fallon WM, et al. Unruptured intracranial aneurysms: natural history, clinical outcome, and risks of surgical and endovascular treatment. Lancet. 2003;362:103–110. doi: 10.1016/s0140-6736(03)13860-3

6. Greving JP, Wermer MJ, Brown RD, Jr., Morita A, Juvela S, Yonekura M, Ishibashi T, Torner JC, Nakayama T, Rinkel GJ, et al. Development of the PHASES score for prediction of risk of rupture of intracranial aneurysms: a pooled analysis of six prospective cohort studies. Lancet Neurol. 2014;13:59–66. doi: 10.1016/S1474-4422(13)70263-1

7. Investigators UJ, Morita A, Kirino T, Hashi K, Aoki N, Fukuhara S, Hashimoto N, Nakayama T, Sakai M, Teramoto A, et al. The natural course of unruptured cerebral aneurysms in a Japanese cohort. N Engl J Med. 2012;366:2474–2482. doi: 10.1056/NEJMoa1113260

8. Byoun HS, Huh W, Oh CW, Bang JS, Hwang G, Kwon OK. Natural History of Unruptured Intracranial Aneurysms : A Retrospective Single Center Analysis. jkns. 2016;59:11–16. doi: 10.3340/jkns.2016.59.1.11

9. Ujiie H, Tachibana H, Hiramatsu O, Hazel AL, Matsumoto T, Ogasawara Y, Nakajima H, Hori T, Takakura K, Kajiya F. Effects of size and shape (aspect ratio) on the hemodynamics of saccular aneurysms: a possible index for surgical treatment of intracranial aneurysms. Neurosurgery. 1999;45:119-129; discussion 129-130. doi: 10.1097/00006123-199907000-00028

10. Tremmel M, Dhar S, Levy EI, Mocco J, Meng H. Influence of intracranial aneurysm-to-parent vessel size ratio on hemodynamics and implication for rupture: results from a virtual experimental study. Neurosurgery. 2009;64:622–630; discussion 630-621. doi: 10.1227/01.Neu.0000341529.11231.69

11. Zhou G, Zhu Y, Yin Y, Su M, Li M. Association of wall shear stress with intracranial aneurysm rupture: systematic review and meta-analysis. Sci Rep. 2017;7:5331. doi: 10.1038/s41598-017-05886-w

12. Cebral JR, Mut F, Weir J, Putman C. Quantitative characterization of the hemodynamic environment in ruptured and unruptured brain aneurysms. AJNR Am J Neuroradiol. 2011;32:145–151. doi: 10.3174/ajnr.A2419

13. Hackenberg KAM, Hänggi D, Etminan N. Unruptured Intracranial Aneurysms. Stroke. 2018;49:2268–2275. doi: doi:10.1161/STROKEAHA.118.021030

14. Dhar S, Tremmel M, Mocco J, Kim M, Yamamoto J, Siddiqui AH, Hopkins LN, Meng H. Morphology parameters for intracranial aneurysm rupture risk assessment. Neurosurgery. 2008;63:185-196; discussion 196-187. doi: 10.1227/01.NEU.0000316847.64140.81

15. Nader-Sepahi A, Casimiro M, Sen J, Kitchen ND. Is aspect ratio a reliable predictor of intracranial aneurysm rupture? Neurosurgery. 2004;54:1343-1347; discussion 1347-1348. doi: 10.1227/01.neu.0000124482.03676.8b

16. Ujiie H, Tamano Y, Sasaki K, Hori T. Is the aspect ratio a reliable index for predicting the rupture of a saccular aneurysm? Neurosurgery. 2001;48:495-502; discussion 502-493. doi: 10.1097/00006123-200103000-00007

17. Lee KS, Zhang JJY, Alalade AF, Vine R, Lanzino G, Park N, Roberts G, Gurusinghe NT. Radiological surveillance of small unruptured intracranial aneurysms: a systematic review, meta-analysis, and meta-regression of 8428 aneurysms. Neurosurg Rev. 2021;44:2013–2023. doi: 10.1007/s10143-020-01420-1

18. Rahman M, Ogilvy CS, Zipfel GJ, Derdeyn CP, Siddiqui AH, Bulsara KR, Kim LJ, Riina HA, Mocco J, Hoh BL. Unruptured cerebral aneurysms do not shrink when they rupture: multicenter collaborative aneurysm study group. Neurosurgery. 2011;68:155-160; discussion 160-151. doi: 10.1227/NEU.0b013e3181ff357c

19. Staarmann B, Smith M, Prestigiacomo CJ. Shear stress and aneurysms: a review. Neurosurg Focus. 2019;47:E2. doi: 10.3171/2019.4.FOCUS19225

20. Han KS, Lee SH, Ryu HU, Park SH, Chung GH, Cho YI, Jeong SK. Direct Assessment of Wall Shear Stress by Signal Intensity Gradient from Time-of-Flight Magnetic Resonance Angiography. Biomed Res Int. 2017;2017:7087086. doi: 10.1155/2017/7087086

21. Lee WJ, Jeong SK, Han KS, Lee SH, Ryu YJ, Sohn CH, Jung KH. Impact of Endothelial Shear Stress on the Bilateral Progression of Unilateral Moyamoya Disease. Stroke. 2020;51:775–783. doi: 10.1161/STROKEAHA.119.028117

22. Lee CH, Lee SH, Cho YI, Jeong SK. Association of Carotid Artery Endothelial Signal Intensity Gradient with Unilateral Large Artery Ischemic Stroke. Cerebrovasc Dis. 2021;50:270–278. doi: 10.1159/000514141

23. McKinney AM. Artifacts of the Craniocervical Arterial System on MRI. In: Atlas of Normal Imaging Variations of the Brain, Skull, and Craniocervical Vasculature. Cham: Springer International Publishing; 2017:1261–1291.

24. Hoh BL, Sistrom CL, Firment CS, Fautheree GL, Velat GJ, Whiting JH, Reavey-Cantwell JF, Lewis SB. Bottleneck factor and height-width ratio: association with ruptured aneurysms in patients with multiple cerebral aneurysms. Neurosurgery. 2007;61:716–722; discussion 722-713. doi: 10.1227/01.NEU.0000298899.77097.BF

25. Baharoglu MI, Schirmer CM, Hoit DA, Gao BL, Malek AM. Aneurysm inflow-angle as a discriminant for rupture in sidewall cerebral aneurysms: morphometric and computational fluid dynamic analysis. Stroke. 2010;41:1423–1430. doi: 10.1161/STROKEAHA.109.570770

26. Shojima M, Oshima M, Takagi K, Torii R, Hayakawa M, Katada K, Morita A, Kirino T. Magnitude and role of wall shear stress on cerebral aneurysm: computational fluid dynamic study of 20 middle cerebral artery aneurysms. Stroke. 2004;35:2500–2505. doi: 35/11/2500 [pii] 10.1161/01.STR.0000144648.89172.0f

27. Hanley JA, McNeil BJ. The meaning and use of the area under a receiver operating characteristic (ROC) curve. Radiology. 1982;143:29–36.

28. DeLong ER, DeLong DM, Clarke-Pearson DL. Comparing the Areas under Two or More Correlated Receiver Operating Characteristic Curves: A Nonparametric Approach. Biometrics. 1988;44:837–845. doi: 10.2307/2531595

29. Jung K-H. New Pathophysiological Considerations on Cerebral Aneurysms. Neurointervention. 2018;13:73–83. doi: 10.5469/neuroint.2018.01011

30. Zhang Y, Jing L, Zhang Y, Liu J, Yang X. Low wall shear stress is associated with the rupture of intracranial aneurysm with known rupture point: case report and literature review. BMC Neurol. 2016;16:231. doi: 10.1186/s12883-016-0759-0

31. Thompson BG, Brown RD, Jr., Amin-Hanjani S, Broderick JP, Cockroft KM, Connolly ES, Jr., Duckwiler GR, Harris CC, Howard VJ, Johnston SC, et al. Guidelines for the Management of Patients With Unruptured Intracranial Aneurysms: A Guideline for Healthcare Professionals From the American Heart Association/American Stroke Association. Stroke. 2015;46:2368–2400. doi: 10.1161/str.0000000000000070

32. Tarumi T, Zhang R. Cerebral blood flow in normal aging adults: cardiovascular determinants, clinical implications, and aerobic fitness. J Neurochem. 2018;144:595–608. doi: 10.1111/jnc.14234

33. Torbey MT, Hauser TK, Bhardwaj A, Williams MA, Ulatowski JA, Mirski MA, Razumovsky AY. Effect of age on cerebral blood flow velocity and incidence of vasospasm after aneurysmal subarachnoid hemorrhage. Stroke. 2001;32:2005–2011. doi: 10.1161/hs0901.094622

34. Duan Z, Li Y, Guan S, Ma C, Han Y, Ren X, Wei L, Li W, Lou J, Yang Z. Morphological parameters and anatomical locations associated with rupture status of small intracranial aneurysms. Sci Rep. 2018;8:6440. doi: 10.1038/s41598-018-24732-1

35. Sonobe M, Yamazaki T, Yonekura M, Kikuchi H. Small unruptured intracranial aneurysm verification study: SUAVe study, Japan. Stroke. 2010;41:1969–1977. doi: 10.1161/STROKEAHA.110.585059

